# Appraisal of Gene Expression-Based Classifiers for Neuropsychiatric Disorders: A Meta-Regression

**DOI:** 10.1101/2024.10.02.24314719

**Authors:** Ali Razavi, Brittany Arensman, Eric J. Barnett, Leo A. Lee, Stephen V. Faraone, Stephen J. Glatt, Jonathan L. Hess

## Abstract

A substantial body of research examines the potential of gene-expression-based biomarkers for diagnosing and selecting treatments for neuropsychiatric disorders, yet no clear consensus has been reached regarding the influence of controllable factors such as study design and model selection on the performance of gene-expression-based classifiers. To investigate study characteristics and methodologies that influence the accuracy of studies using transcriptomics to classify neuropsychiatric disorders, we conducted a literature review and meta-regression of relevant studies. We extracted several characteristics from each study, including the number of samples in a training dataset, approach for model validation, and classification model. Using univariate and multi-variate mixed-effect meta-regression analyses, we estimated the association between these study characteristics and reported classification accuracies. Machine Learning (ML) models accounted for 55% of all models, Deep Learning (DL) models accounted for 20% and variations of Logistic Regression models making up the remaining 25%. Support vector machine(SVM) was the most common model type (17%).The use of withheld test samples (56%) was the most frequent approach for validating performance of classification models. We found significant associations between reported accuracies and study-rated bias risk, model type, class ratio, and validation approach. Overall, this review provides helpful insights into study characteristics that significantly influence classification accuracies and emphasizes the importance of prudent methodologies for training and evaluating classification models to mitigate biased accuracy estimates.

## INTRODUCTION

Over the past two decades, researchers have explored biomarkers associated with neuropsychiatric disorders, aiming to determine their capacity for accurate and objective identification of these disorders. These biomarkers have been evaluated from many modalities including genetics, transcriptomics, proteomics, epigenetics, metabolomics, and brain structural and functional activity. Due to the complexity and overlapping of features in neuropsychiatric disorders, developing algorithms for diagnosing patients or predicting treatment outcomes has been a significant challenge (Cornblath et al., 2019). Machine learning (ML) and Deep Learning (DL) methods have emerged as powerful tools for gaining insights into complex biology and disease, demonstrating considerable potential in uncovering useful biomarker patterns.

Significant strides have been achieved in transcriptomic technology, expanding the number and diversity of RNAs that can be profiled in cells and tissues. This progress, coupled with substantial advances in genomics, has paved the way for in-depth explorations of transcript variants and splicing isoforms. These developments have prompted further investigations into the harnessing of the potential of RNA expression states as valuable diagnostic or therapeutic biomarkers. Many studies that investigated the diagnostic utility of gene expression-based biomarkers for neuropsychiatric disorders have used classification methods that range from conventional regression-based models, like logistic regression, to ML-based methods such as support vector machine or neural networks. These studies have used a wide array of study designs and procedures to evaluate model performance. This procedural variability among studies poses an opportunity to investigate how study characteristics influence model performance. Earlier reviews of classification studies have found that the size of the training sample and procedures used for assessing model performance are among the most significant predictors of classification accuracy for genomics (Barnett et al., 2024) and brain structural imaging (Zhang-James et al., 2023; Vabalas et al., 2019). To date, neuropsychiatric studies that evaluated classification performance for ML-based methods have commonly relied upon smaller sample sizes, falling below the recommended sample size (Vabalas et al., 2019) criteria for modeling complex tasks. This is particularly pertinent for genomics datasets, where the number of features often surpasses the number of samples by several orders of magnitude. Ascertaining tens-to-hundreds of thousands of samples for an individual classification study is generally neither practical nor feasible. Therefore, when investigators apply ML to smaller-sized datasets, it is crucial to adhere to standard procedures to ensure to prudently use the methods and evaluate them, thus mitigating the risk of bias (Vabalas et al., 2019). For example, it has been shown that k-fold cross-validation tends to strongly bias performance estimates, but nested cross-validation and designs that use dedicated training and testing split validation methods yield more accurate and less overfitted results (Vabalas et al., 2019; Quinn & Hess et al., 2024; Zhang-James et al., 2023).

Several reviews highlight the potential for using blood-based gene expression-based biomarkers to individualize treatments for neuropsychiatric disorders, including depression (Mariani et al., 2021), schizophrenia (Mamdani et al., 2013), and Alzheimer’s Disease (Teunissen et al., 2022). The potential of blood-based biomarkers to reveal molecular pathways contributing to the etiology of neuropsychiatric disorders has also been reviewed, accompanied by recommendations for applying their clinical application (van de Leemput, Glatt, & Tsuang, 2016). Several studies have examined the utility of blood-based gene-expression and other ‘omics profiles to serve as useful proxies for the brain. The overarching consensus from these studies is that the expression levels of genes highly expressed in both the brain and blood exhibit a moderate correlation across these tissues (Tylee et al., 2013). These findings have been expanded upon by other studies, revealing that several gene co-expression networks in the human brain are significantly and strongly preserved in the blood (Cai et al., 2010; Hess et al., 2016). This work highlights the considerable potential of blood-based gene-expression to serve as a useful proxy of the human brain. Recent advancements in transcriptome imputation methodologies, such as *BrainGENIE, B-GEX, and TEEBoT*, have further enhanced this potential by enabling gene expression profiles in the brain to be predicted based on gene expression profiles directly measured in the blood (Hess et al., 2023; Basu & Wang et al., 2021; Xu et al., 2020). The present study is a literature review and meta-regression of the gene expression-based classification studies centered on neuropsychiatric disorders aiming to investigate the study characteristics and methodologies that significantly influence the accuracy of the classification models using transcriptomics to predict neuropsychiatric disorders.

## METHODS

### Literature Search

We conducted a literature search in PubMed to identify original research studies that evaluated diagnostic- or treatment-response based classifiers based on traditional statistical models (such as logistic regression) or supervised ML algorithms (such as *k*-nearest neighbors, linear discriminant analysis, support vector machine, random forest, neural networks, XGBoost, and Naïve Bayes, to name a few) in the context of neuropsychiatric disorders. The scope of our review focused on studies that used gene expression exclusively or in combination with other data modalities as input into classification models. We searched PubMed and Medline using the following keywords: (Alzheimer’s disease OR mild cognitive impairment OR attention-deficit/hyperactivity disorder OR schizophrenia OR depression OR bipolar disorder OR autism OR post-traumatic stress disorder OR suicide*) AND (gene expression OR transcript*) AND (classification OR machine learning OR support vector machine OR random forest OR neural network OR decision tree OR naive bayes OR XGBoost OR linear discriminant analysis OR nearest neighbor OR deep learning OR elastic net OR lasso OR gradient boosting OR autoencoder OR long-short term memory OR gated recurrent units) AND (AUC or accuracy OR area under the curve). Our literature search yielded 260 studies and their full texts were reviewed. Studies that did not report accuracy or area under the receiver operating characteristic curve (AUC) measures were excluded. Studies that reported diagnostic accuracy of gene expression-based biomarkers solely *via* a ROC analysis without any implementation of ML or regression-based classifiers were excluded. A total of 198 studies were retained for our review.

### Data Extraction

The full-text articles were reviewed and the following study features were recorded for our analysis: the number of samples in training dataset per class, number of samples in the validation set per class (if available), approach for model validation (none, *k*-fold cross-validation (CV), leave-one-out cross-validation (LOOCV), divided training/test sets), choice of classification model, and performance metrics of classification model (*i.e.,* AUC, sensitivity, specificity, or overall accuracy). If studies reported multiple accuracies for different parametrizations of the same classification model (*e.g.,* multiple SVMs trained using the top 5, 10, … *k* differentially expressed genes), then the best-performing model was recorded for our analysis. As some studies did not report how many subjects were randomly assigned to training and validation sets, we approximated these numbers from the reported percentage of subjects included in each set (*e.g.,* 80% in training set and 20% in test set).

We evaluated studies for potential bias based on the following conditions:

1. Feature selection was performed exclusively on the samples designated for model training.
2. The training sample comprised a balanced set of classes or corrections were made to address class imbalance.
3. Performance of the classification model was evaluated on a set of data treated separate from the training data

Studies for which all three conditions were satisfied were assigned a “low likelihood” risk for potential bias. Studies for which one or more conditions were not satisfied or if unclear were assigned “probable/high likelihood” risk for potential bias.

A total of 92 studies were retrieved and 532 models were extracted. Ten of the models were excluded due to not reporting classification performance in terms of AUC or overall accuracy (n=2) or failing to report sample size of the training set (n=9), leaving 521 total models from 91 studies that were used in the analyses.

### Statistical Analysis

We performed univariate linear mixed-effect regression analyses to estimate the association between each study characteristic and reported classification performance. In our univariate regression analyses, we specified each study characteristic as an independent variable and reported accuracy as the dependent variable. We corrected *p*-values obtained by our univariate models using the Benjamini-Hochberg false discovery rate (FDR) procedure. To gain additional insight into the joint effect of study characteristics on reported accuracy, we conducted a meta-regression in which we modeled all study characteristics as predictors of reported classification performance *via* a linear mixed-effect regression model. All mixed-effect regression models included a random-effect term for study to account for the non-independence among study observations. Standard errors were approximated using the Satterthwaite approach. All statistical analyses were performed in *R* (v.4.2.1) and mixed-effect regression models were fitted using the *lmerTest* package.

### Secondary Analysis of Reported Accuracies in Relation to Tissue Source of Biomarkers

It is plausible that biomarkers obtained from *postmortem* brain tissue analyses exhibit greater ontological relevance to psychiatric disorders than peripherally accessible RNAs, potentially influencing classification performance. Additionally, we suspected that tissue concordance (*i.e.,* using either exclusively peripheral blood or *postmortem* brain for both model training and validation) or discordance (*i.e*., transition models between brain and blood samples for model training and validation) between datasets might impact classification accuracy. We conducted a secondary analysis on a subset of our entire study pool, comprising 51 unique studies and 292 classification models. These studies evaluated the performance of classification models on completely withheld samples, independent from those used for model training. Among these models, 233 were trained on gene expression profiles from *ex vivo* peripheral blood samples and tested on independent *ex vivo* peripheral blood data. Additionally, two studies trained models on gene expression profiles observed in *ex vivo* blood samples and validated them using data obtained from *postmortem* brain tissue. Nine studies trained models using gene expression profiles observed in *postmortem* brain tissue and validated them using data from *ex vivo* peripheral blood. Finally, 37 models were trained on gene expression profiles observed in *postmortem* brain and validated in independent *postmortem* brain samples. We compared reported classification accuracies between these four groups of studies using a mixed-effect type III ANOVA using a unique study identifier as a random-effect term to adjust for repeated measurements. A significance threshold of *p<*0.05 was used for this *post-hoc* analysis.

## RESULTS

### Overview of Study Characteristics

Our literature search uncovered 91 eligible studies, as detailed in the PRISMA diagram provided in **Supplementary Figure 1**. A complete collection of study data is provided in **Supplementary File 1**. These studies reported classification accuracies for 521 models for 13 neuropsychiatric disorders, including: autism spectrum disorder, Alzheimer’s disease, mild cognitive impairment, schizophrenia, bipolar disorder, psychosis, obsessive-compulsive disorder, attention-deficit/hyperactivity disorder, methamphetamine-associated psychosis, suicide, major depressive disorder, post-traumatic stress disorder, and Parkinson’s disease.

**Figure 1a** summarizes the number and type of classification models reported per disorder. ML-based algorithms were used in 289 (55%) of reported classification models, DL-based models in 105 (20%), logistic regression models accounted for 85 of the models (16%) and regularized logistic regression models accounted for 42 of the models (9%). Support vector machine (SVM) was the most frequently used classification model among the studies in our review, which accounted for 17% or 86 of the 521 models. A low potential risk of bias was found for 74 out of the 91 studies; 17 studies showed probable/high potential risk of bias. Classification models for Alzheimer’s disease and mild cognitive impairment accounted for 332 (63%) of all reported classifiers. The most frequent validation approach for evaluating classification model performance was *via* withheld test sample (292 models), followed by k-fold cross-validation (189 models), and LOOCV (21 models). In 19 models, classification performance was reported based solely on a training sample without the use of cross-validation or an independent test sample. An overview of the number of studies that contributed to each disorder is provided in **Figure 1b**. A complete collection of study data is provided in **Supplementary File 1**.

**Figure 1.**
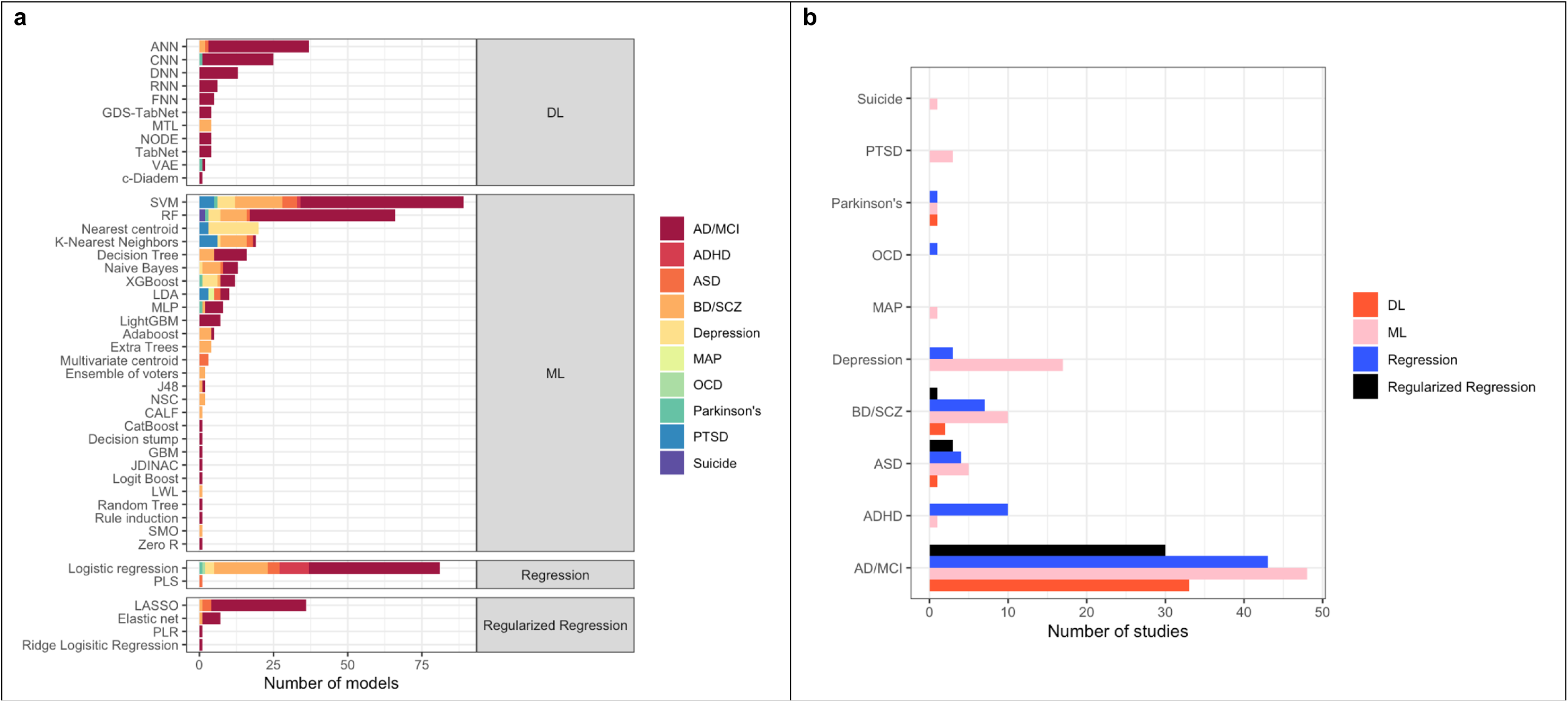
Graphical overview of the gene-expression based classification studies included in our meta-regression. **(a)** A bar-plot showing the number of classification models used for each of the main neuropsychiatric disorders represented among studies in our meta-regression dataset. (**b**) The number of studies featured per classification model type for each of the neuropsychiatric disorders represented in our dataset. Note: To enhance visualization clarity, we grouped Alzheimer’s disease (AD) and mild cognitive impairment (MCI)-focused classifiers into a single diagnostic label (“AD/MCI”). Similarly, we grouped schizophrenia (SCZ), bipolar disorder (BD), and psychosis into a single diagnostic label (“SCZ/Psychosis/BD”). Other abbreviations: attention-deficit/hyperactivity disorder (ADHD), autism spectrum disorder (ASD), methamphetamine-associated psychosis (MAP), major depressive disorder (MDD), Parkinson’s disease (PD), post-traumatic stress disorder (PTSD).

There was a significant difference in reported classification performance across diagnostic groups when accounting for bias level and sample size as illustrated in **Figure 2** (p= 0.029). The mean accuracy for all models was 0.72 (SD= 0.16).

**Figure 2.**
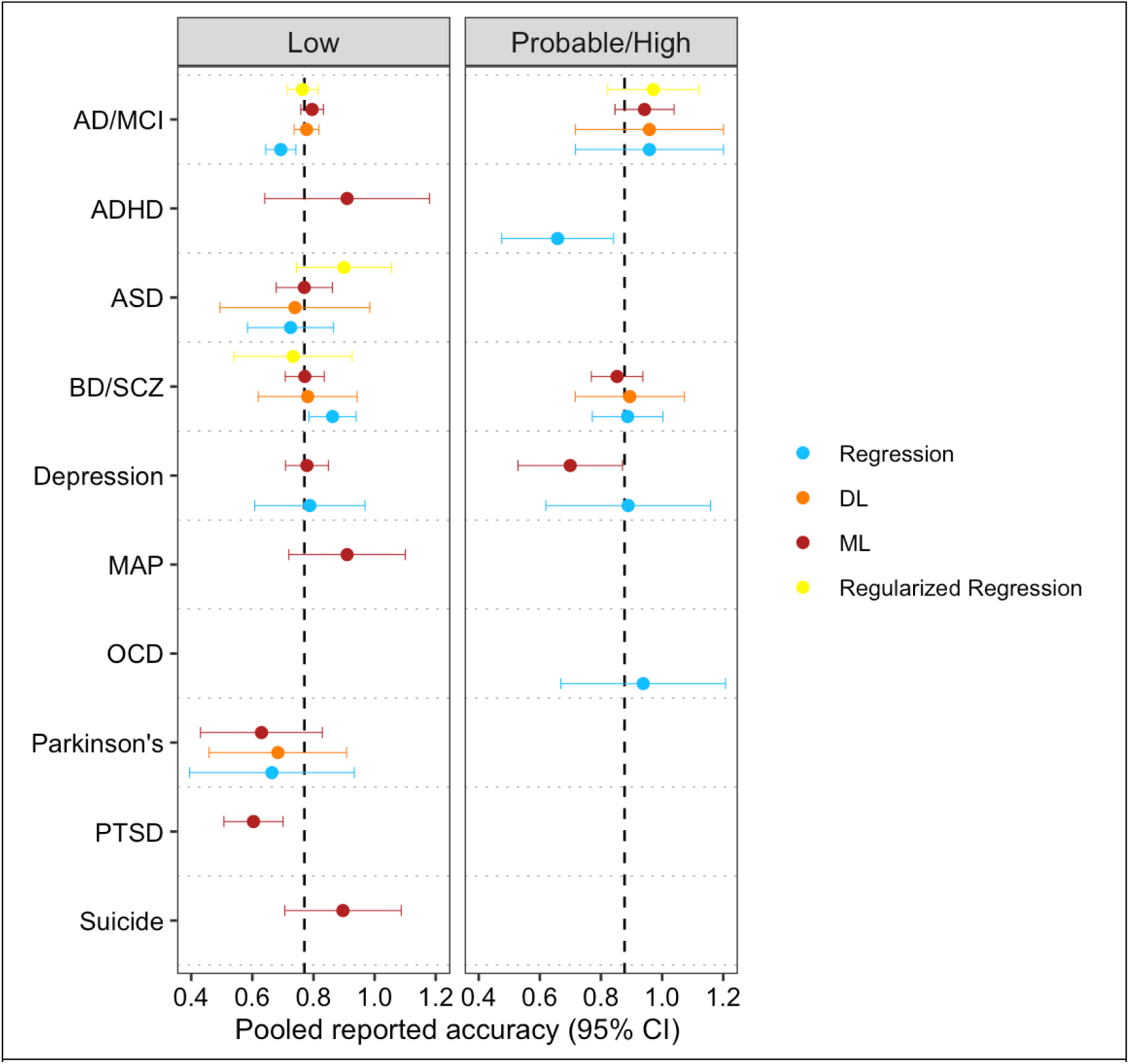
Forest plot showing the pooled estimated marginal means in reported accuracy for each diagnostic group. Pooled estimates of reported accuracy were obtained using a univariate linear mixed-effect model to adjust for repeated measures from studies. Estimated mean reported accuracies are color coded by type of classification model as follows: regression models appear in blue, machine learning models appear in orange, and neural networks appear in red. Vertical dashed lines designate the average pooled reported accuracy among all models according to study bias rating. There was a significant difference in reported accuracy across diagnostic groups according to mixed-effect type III ANOVA accounting for bias risk and model type (F_9,111.17_ = 2.1663, *p* = 0.029). Some categories do not include points in the figure due to limitations of estimated marginal means calculation for small sample sizes. **Abbreviations**: Alzheimer’s disease (AD), mild cognitive impairment (MCI), attention-deficit/hyperactivity disorder (ADHD), autism spectrum disorder (ASD), bipolar disorder (BD), methamphetamine-associated psychosis (MAP), major depressive disorder (MDD), Parkinson’s disease (PD), post-traumatic stress disorder (PTSD), schizophrenia (SCZ)

### Univariate Mixed-Effect Meta-Regression of Study Characteristics and Reported Accuracy

The results obtained from univariate mixed-effect meta-regression models are summarized in **Figures 3 and 4**. We found a significant association between study-rated bias risk and classification performance (*p*=0.004). Studies rated as having high/unclear bias exhibited higher reported accuracies than studies with low observed bias. Because high risk of bias can inflate reported classification accuracy, we included study-rated bias risk as a covariate in our downstream multivariate meta-regression when assessing the influence of other study characteristics on reported accuracy.

**Figure 3.**
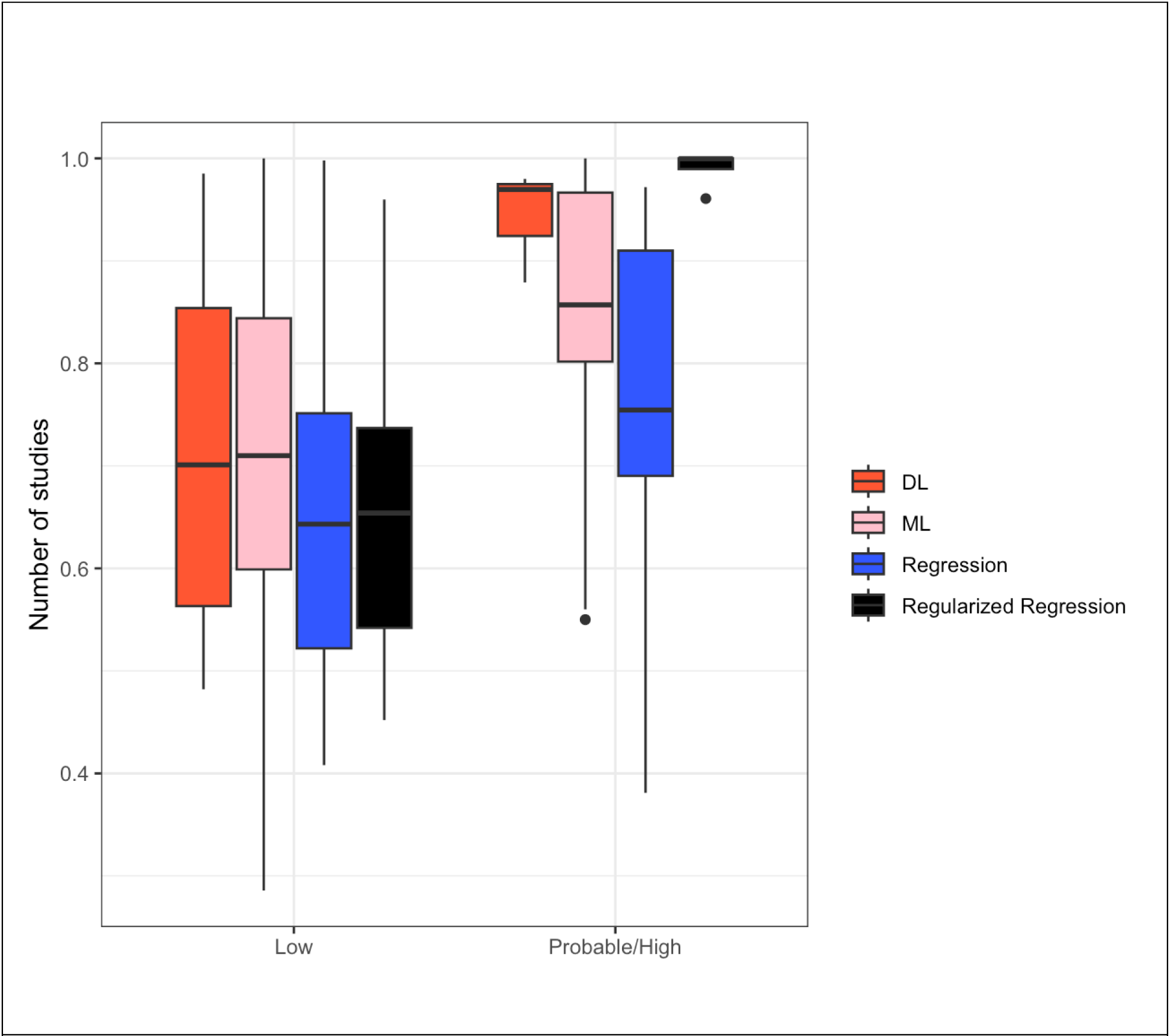
Accuracy of models stratified by their risk of study bias and type of classification model. **Abbreviations**: machine learning (ML), deep learning (DL). A type III mixed-effect ANOVA revealed a significant group-wise difference in reported accuracy between studies with low vs. probable/high bias (F_1, 291.90_= 8.3512, *p* = 0.004143), but no significant difference was observed based on the interaction between bias rating and type of classification model.

Our univariate mixed-effect regression analyses showed a small yet significant association between the size of the training sample and classification accuracy (**Figure 4a**). Notably, these results revealed an inverse relationship: as sample size increased, reported accuracies decreased by a rate of 0.056, 0.038, per unit increase in log_10_-transformed sample size for studies using k-fold CV and LOOCV respectively.

**Figure 4.**
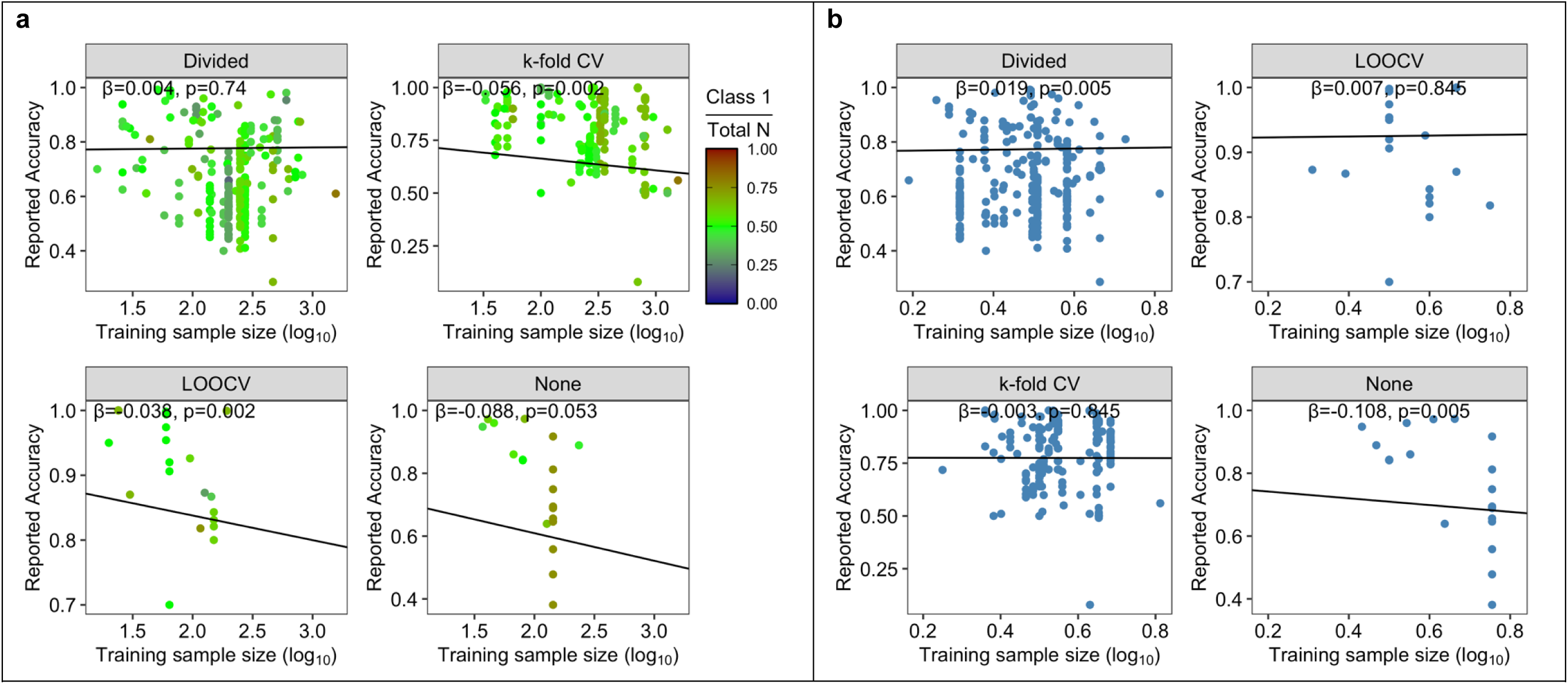
Scatterplots depicting the univariate linear relationship between reported classification model accuracies and (left) the log_10_-transformed total sample size of the training dataset (x-axis), and (right) the proportion of positive classes in the training dataset. Plots were stratified by types of validation procedure used in the study. Each dot represents a single classification model. The regression coefficients and *p*-values from a univariate mixed-effect regression model are provided in each plot, which account for non-independence of the studies in our meta-regression. Black solid lines denote the best-fit relationship between variables based on mixed-effect linear regression.

We found a significant inverse association between classification performance and the ratio of cases to controls within training samples. This association was found among studies that evaluated classification performance within independent test samples (*p*=0.005) and studies where performance was solely evaluated in the training sample (*p*=0.005) (**Figure 4b**). Type of accuracy was not significantly associated with reported accuracy and had no impact on the association between ratio of cases to controls within training samples and accuracy (**Figure 4b**).

As shown in **Supplementary Figure 2**, no significant association was found between year of study and study accuracy (*p*=0.09). However, year of study was significantly correlated with total size of training sample (*p*=4.9 x 10^-10^). All linear models were analyzed for heteroscedasticity through the application of Gamma family Generalized Linear Models with a log link and showed similar statistical significance.

### Multivariate Mixed-Effect Meta-Regression Analysis of Study Characteristics and Reported Accuracy

**Table 1** provides the summary statistics from the multivariate linear mixed-effect Regression model, which was used to estimate the conditional effects of study-level characteristics on reported accuracy of classification models. From an omnibus mixed-effect ANOVA, reported classification accuracy was found to be significantly associated with all six study characteristics. The significant association were found for the total sample size of the training set (F-value=13.96, p=0.0003), type of classification model (F-value=6.10, p=0.0004), class ratio (F-value=7.24, p=0.007), approach used by studies to assess validation of model performance (F-value=7.95, p=6 x 10^-5^), the diagnostic group studied (F-value = 4.19, p=0.0001), and study-rated bias risk (F-value=6.59, p=0.01).

**Table 1.** Summary statistics from a type III mixed-effect analysis of variance (ANOVA) and mixed-effect linear regression models. The ANOVA model was used to estimate the omnibus association of each predictor variable, particularly categorical variables that had 3+ factor levels (*i.e.,* validation type and classified type), on reported mean accuracy of the studies included in our meta-regression. Summary statistics on the right-hand side of the table (i.e., not under the columns designated for the ANOVA statistics) are the regression coefficients indicating the main effects of continuous *or* differences in group means for 2-level categorical values on reported classification accuracies. For categorical variables with 3+ groups, we provide the pair-wise differences in estimated marginal means. In the mixed-effect ANOVA and regression models, a unique study indicator was set as the random-effect variable to adjust for non-independence between model accuracies.

|  | Type III Mixed-effect ANOVA |  |  | Estimated marginal means (and *post-hoc* comparisons) |  |  |  |
| --- | --- | --- | --- | --- | --- | --- | --- |
| Variable | F-value | d.f. | p-value | Beta* | S.E. | t-value | p-value* |
| **Diagnostic group\*** | 4.1857 | 9, 106.42 | **0.0001195** |  |  |  |  |
| AD/MCI vs. MDD |  |  |  | 0.16 | 0.039 | 4.10 | **0.002** |
| AD/MCI vs. PTSD |  |  |  | 0.22 | 0.048 | 4.54 | **0.0009** |
| ASD vs PTSD |  |  |  | 0.178 | 0.554 | 3.221 | **0.0272** |
| BD/SCZ vs. PTSD |  |  |  | 0.152 | 0.050 | 3.02 | **0.0386** |
| **Class ratio** | 7.3524 | 1, 451.03 | **0.0069530** | 0.17 | 0.065 | 2.62 | **0.0089** |
| **Classifier type** | 6.1037 | 3 495.92 | **0.0004400** |  |  |  |  |
| ML vs. Regression |  |  |  | 0.07227 | 0.017 | - 4.157 | **0.0002** |
| DL vs. Regression |  |  |  | 0.07117 | 0.021 | 3.421 | **0.0020** |
| **Bias risk**<br>(Probable/High vs. Low) | 6.5874 | 1 126.63 | **0.0114339** | 0.0743 | 0.029 | 2.547 | **0.0122** |
| **Validation type** | 7.9538 | 3,148.35 | **5.928e-05** |  |  |  |  |
| Divided vs. k-fold CV |  |  |  | -0.00073 | 0.027 | 0.055 | 0.96 |
| Divided vs. LOOCV |  |  |  | -0.1998 | 0.042 | -4.73 | **<10<sup>-22</sup>\*** |
| Divided vs. None |  |  |  | -0.0877 | 0.0515 | -1.703 | 0.1170 |
| k-fold CV vs. LOOCV |  |  |  | -0.1991 | 0.0453 | -4.394 | **0.001** |
| k-fold CV vs. None |  |  |  | -0.0869 | 0.0522 | -1.666 | 0.1170 |
| LOOCV vs. None |  |  |  | 0.112137 | 0.0608 | 1.846 | 0.1170 |
| **Total sample size, log<sub>10</sub>** | 13.958 | 1,139.99 | **0.0002713** | -0.085 | 0.03 | -2.83 | **0.0055** |

**Note:** Degrees of freedom for the mixed-effect regression were estimated using Satterthwaite's method. **Beta\*:** indicates the regression coefficient (slope) of the main effect for continuous variables on reported accuracy, or differences in estimated marginal means of reported accuracy between categorical variables. **p-value\*:** the column title denoting *p*-values indicates that *post-hoc* pairwise comparisons were adjusted for multiple comparisons with Benjamin-Hochberg false-discovery rate procedure. \*\*:values that could not be printed with more precision. **Diagnostic group\*:** As there were a large number of *post-hoc* comparisons with respect to the diagnostic group variable, we present only the statistically significant findings in the table.

Post-hoc pair-wise tests revealed significant differences in classification performance among categorical study characteristics. Studies employing logistic regression-based classifiers exhibited significantly lower classification accuracies compared to those using ML models (p= 0.0002). Logistic regression-based classifiers also exhibited significantly lower classification accuracies compared to DL models (p= 0.002). In addition, studies using LOOCV exhibited significantly higher classification accuracies compared to studies that utilized withheld independent test sets (p <10^-22^), as well as studies that used k-fold CV (p=0.001). Furthermore, we found that studies that reported classification accuracies for models tasked with detecting AD/MCI were significantly higher than models tasked with detecting PTSD (p=0.0009). Studies that reported classification accuracies for models tasked with detecting AD/MCI were significantly higher than models tasked with detecting MDD (p= 0.002). Studies that reported classification accuracies for ASD were significantly higher than PTSD models (p= 0.027). Studies that reported classification accuracies for BD/SCZ were significantly higher than models focused on PTSD (p= 0.039).

### Relationship between Classification Performance and Tissue Source of Biomarkers

We found no significant difference in reported classification accuracies between studies that were trained on blood or brain data and tested on blood or brain data, suggesting that biomarker sourced from *postmortem* brain, despite their potential ontological relevance for psychiatric disorders, does not improve classification performance as compared to direct blood data (**Figure 5**). Moreover, whether the training and validation sets were acquired from the same tissue-type (either blood or brain tissue), did not significantly impact classification accuracies.

**Figure 5.**
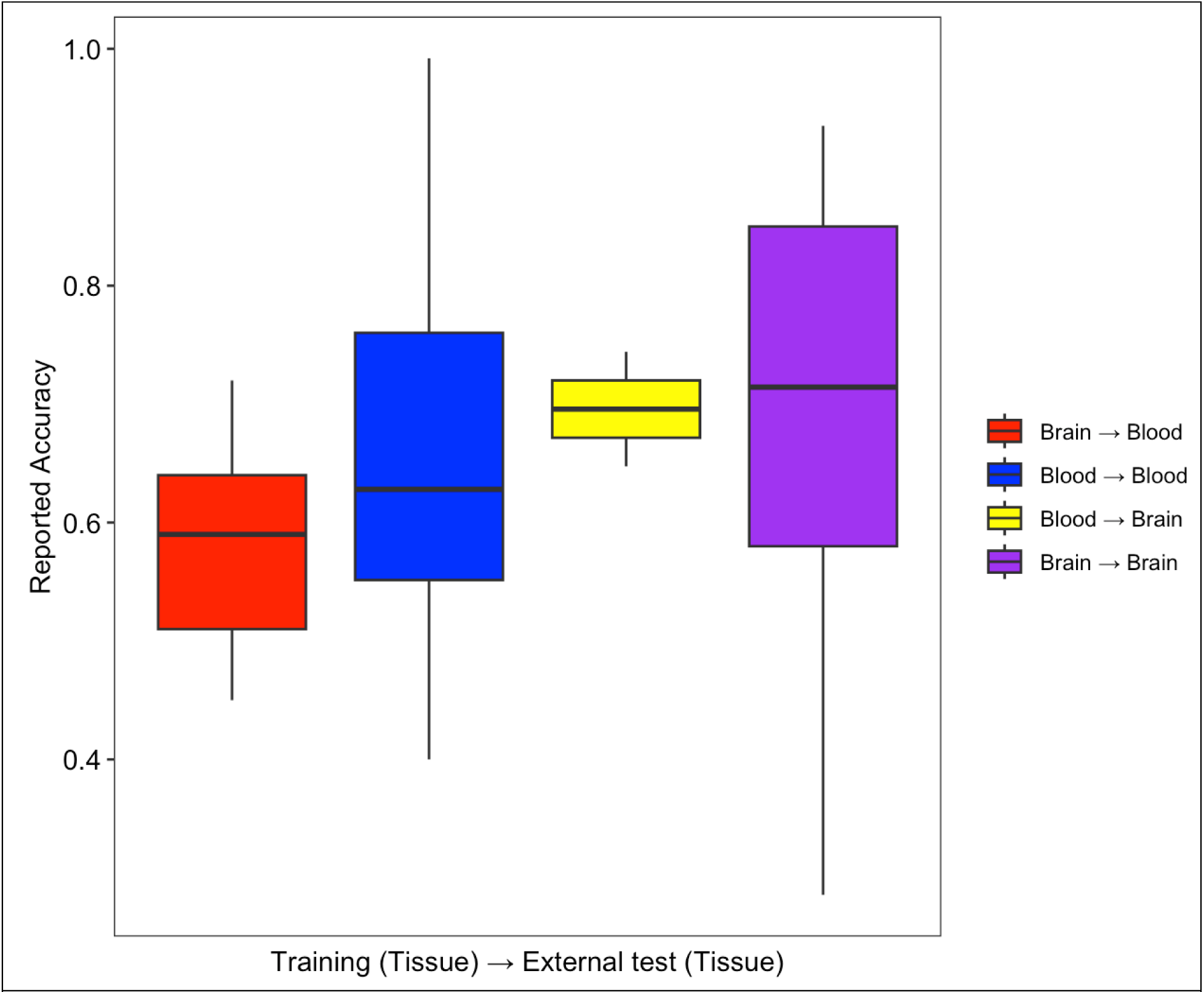
Reported accuracy per training and validation pair in studies. No statistically significant difference in mean reported accuracy was found across the four pairs based on mixed-effect type III ANOVA (F_3,168.02_ = 0.2681, *p* = 0.8483).

## DISCUSSION

This review summarizes the main model and study design variables impacting gene-expression classifier performance in neuropsychiatric disorders. We found that these factors significantly influence the reported performance of gene expression-based classifiers, underscoring the importance of careful interpretation. If these factors are not thoroughly considered during key stages of study design, including model training and validation, the reported performance of gene expression-based classifiers may be either over- or under-estimated, which can lead to misleading interpretations of the utility of these models by investigators. We highlight recommendations from the literature on best practices for mitigating sources of bias, providing guidance for investigators to enhance the robustness and performance of their models (Vabalas, et al., 2019; Zhang-James, et al., 2023; Barnett et al., 2024).

A prevailing challenge with applying classification models to high-dimensional data is the establishment of reliable comparative benchmarks. This can be attributed to the heterogeneity of psychiatric diagnoses, novelty of modeling approaches for diagnosis, and lack of standardized reference performances. Our review provides estimates of the mean and range of reported classification accuracies of gene-expression-based classifiers for specific neuropsychiatric disorders, offering a useful reference point for benchmarking performance in future classification studies. Considering that there was an uneven representation of models in our meta-regression, it is imperative to use caution when extrapolating the estimates and interpreting how the results can be applied.

Our analyses found several trends that should be useful for designing future studies. Firstly, increasing sample size predicted decreasing accuracy with different types of validation methodologies. While this finding may appear counterintuitive, it has been documented in prior meta-analyses of neuropsychiatric biomarker studies (Vabalas, et al., 2019; Zhang-James, et al., 2023). It is likely that studies with smaller sample sizes are more vulnerable to overfitting, resulting in overestimation of model performance. Additionally, variation in sample ascertainment between small and large studies might influence the homogeneity of study groups, potentially influencing estimates of model performance. Inflated estimates from small studies may not be replicable in larger studies that encompass more representative samples capturing the full range of heterogeneity of the disorders of interest. The technical constraints due to small sample size, along with the considerable pathophysiological and clinical heterogeneity inherent in neuropsychiatric disorders, presents challenges for generalizing classification models in future unseen data. These findings underscore the importance of high-quality datasets generated from large, diverse cohorts representative of the population.

We found a small, non-significant trend toward decreasing accuracy among reported classification models over the years, a phenomenon which may influenced by many factors (Rajput, et al., 2022: Guo, Y. et al., 2010). This has been the case in a number of meta-analyses focused on high-dimensional data (Zhang-James, Y., et al., 2023; Xiong, S., et al., 2023). A potential source of model statistic changes over time is increased awareness on the issue of overfitting and model bias. Improved practices that limit study bias such as using standardized approaches for data pre-processing and algorithm deployment may also reduce the risk for over-fitting and over-estimation of classification performance. Choosing a model that exhibits low bias and is the right fit for the data structure is key for improving performance and generalizability of classification models. Optimizing the amount of signal captured by models while steering models away from overfitting requires careful practices to be adopted at various levels of the analysis, including feature selection and/or feature engineering, model design, training, and validation. Additionally, we cannot exclude the potential influence of bias towards better-performing models over the years—commonly known as the “file drawer effect” (Nagarajan et al., 2017). This phenomenon, characterized by the likelihood of positive results being published, may contribute to the observed skewed outcomes over the years.

When dealing with small, cross-validation procedures such as LOOCV and k-fold CV are commonly used for assessing model performance, which allows all available data to be used for training and assessing classifiers. However, the literature suggests that cross-validation yields overly optimistic estimates of model performance compared to studies that employ divided training/test set-ups, especially if the datasets are small and exhibit high variability between the subsets of data for training and testing in each fold, or if there are imbalances between classes in the dataset (Tabe-Bordbar, S., 2018). Over-estimation of model performance can worsen when proper practices are not adhered to in order to prevent data leakage across folds as has been documented for genomic data (Barnett et al., 2024). Leakage of information between subsets of data used for training and validation can happen in numerous ways. One of the most common missteps that lead to data leakage involves applying feature selection, scaling, or transformations—such as normalization, dimensionality reduction, and oversampling—across the entire dataset prior to splitting the data into subsets for cross-validation or dividing datasets into dedicated training, validation, and/or test sets. When these techniques are applied on the complete dataset rather than solely on the training sample, information can inadvertently leak to the validation and/or test data, leading to inflated estimates of model accuracy. This phenomenon was evident in our meta-regression results, where studies exhibiting high bias through improper practices such as breaking the separation of training and validation subsets, tended to overestimate the performance of their models.

We found that ML models, particularly SVM, were the most common type of classification model employed among studies included in our meta-analysis. The popularity of SVM among studies employing gene-expression based classification methods might be attributed to the relative ease of use of such models, ability to capture higher-order interactions between features that may not be known *a priori*, and the availability of explainability methods. Our meta-regression found that the type of classification model selected by studies had a significant influence on reported classification accuracy. Studies using conventional regression-based methods yielded lower classification accuracies than studies using more complex ML and DL models. This makes sense given that, compared with regression, the ML and DL models can model more complex relationships between features and outcomes and may better model the ground truth of psychiatric disorders which, themselves, have complex biology.

Blood, being a readily accessible source of putative biomarkers, offers practical benefits compared to the challenges associated with collecting brain tissue. Brain tissue is not readily accessible in most clinical settings. Interestingly, our results did not reveal a statistically significant difference among classification models trained and validated on these tissues. This result could be partially due to uneven sample size distribution across groups: 232 models were trained and validated on blood, two were trained on blood and validated on brain tissue, nine were trained on brain and validated on blood, and 37 were trained and validated on brain.

Nevertheless, the result suggests that classification tasks are not particularly tissue-specific in training and validation, bringing into question the idea that tissue-specific or cross-tissue analyses will necessarily lead to improved classification accuracies.

There are advanced modeling methodologies holding potential promise that were not used in the pool of investigated studies. It is possible that models currently not utilized in neuropsychiatric gene expression analyses may perform better than current models (Wolpert & Macready, 1997).

### Conclusions

We identified many cases of high study bias leading to either over-fitting or over-estimations of model performance, underscoring the critical importance of adhering to guidelines for the prudent use of ML- or regression-based classification models throughout the entire process, including study design, implementation, and reporting (Quinn, TP., et al., 2024). Furthermore, the findings from our meta-regression show that studies with relatively small sample sizes produced inflated estimates of model performance, underscoring the importance of large, representative datasets. A significant portion of variability in performance of classification models observed across studies can be attributed to the type of classification model selected, the proportion of cases in the training dataset, study-rated bias risk, and approach for validating classification models. Notably, studies that employed ML- and DL-based models exhibited significantly better classification accuracies than regression-based models, even when controlling for other study factors such as sample size and bias risk. In addition, studies with larger numbers of samples in their training dataset tended to yield lower estimates of model performance, particularly for studies that employed k-fold CV or LOOCV to evaluate model performance. This may reflect that studies with relatively few samples for training and evaluating models are susceptible to over-estimating model performance. Conversely, studies employing CV as the primary method for evaluating model performance may better capture patterns capable of generalizing to future unseen data. Based on the findings from our meta-regression, greater prioritization should be given to studies employing large sample sizes and robust bias control. Leveraging larger, diverse datasets in tandem with rigorous ML techniques holds the potential to substantially enhance the generalizability of gene-expression based classifiers tailored for neuropsychiatric disorders.

## FUNDING ACKNOWLEDGEMENTS

**JLH** is supported by grants from the U.S. National Institutes of Health (R21MH126494, R01NS128535), the Central New York Community Fund, and the NARSAD: The Brain & Behavior Research Foundation (2020 Young Investigator Award). **SJG** is supported by grants from the U.S. National Institutes of Health (R01AG064955 and R21MH126494), the U.S. National Science Foundation, the Sidney R. Baer, Jr. Foundation, and NARSAD: The Brain & Behavior Research Foundation. **SVF** is supported by the European Union’s Horizon 2020 research and innovation programme under grant agreement 965381; NIH/NIMH grants U01AR076092, R01MH116037, 1R01NS128535, R01MH131685, 1R01MH130899, U01MH135970, and Supernus Pharmaceuticals. His continuing medical education programs are supported by The Upstate Foundation, Corium Pharmaceuticals, Tris Pharmaceuticals and Supernus Pharmaceuticals.

## DATA AVAILABILITY STATEMENT

The data used in this paper were obtained from published research papers and repositories referenced throughout this manuscript. Specific details in regards to the datasets and studies investigated can be found in Supplementary File 1.

