## Supplementary Figures for "Appraisal of Gene Expression-Based Classifiers for Neuropsychiatric Disorders: A Meta-Regression"

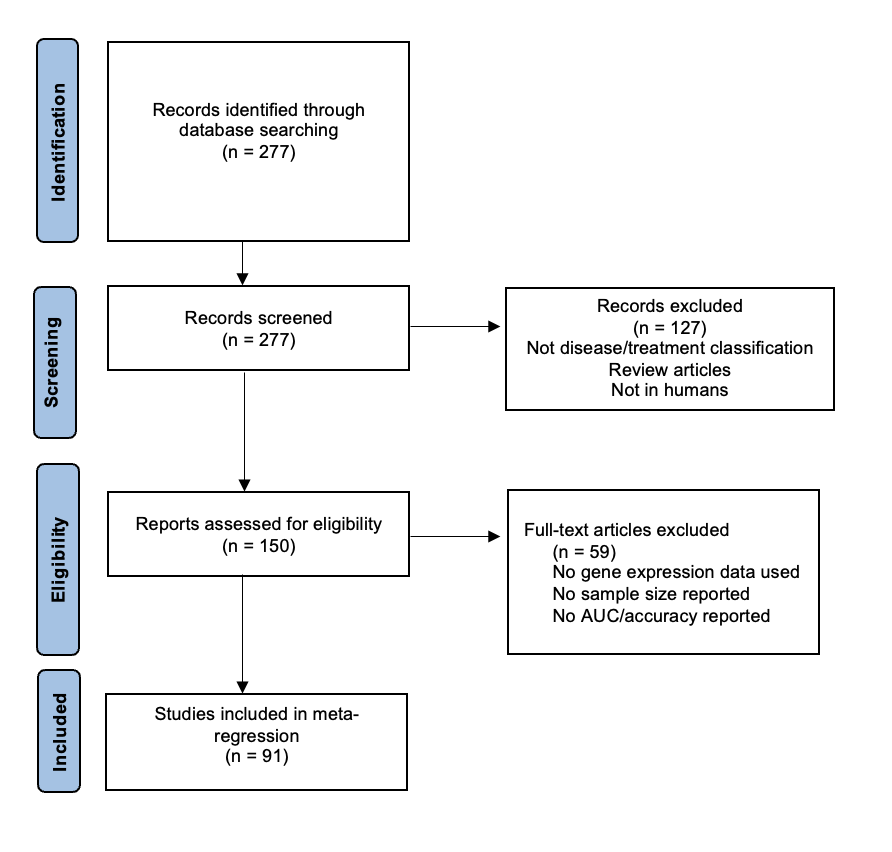

**Supplementary Figure1.** PRISMA diagram of the current study starting from identification/database search. A total of 277 studies were identified using the search criteria outlined in the methods section. Furthermore, 127 studies were excluded due to not having a disease/treatment classification, being review articles or development with non-human data. At the eligibility stage, 59 studies were excluded due to the lack of gene expression data, not reporting sample size and not reporting AUC and accuracy. This process left this study with a total of 91 eligible studies, many of which had multiple reported models.

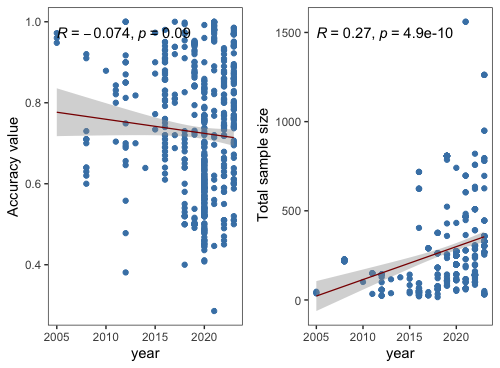

**Supplementary Figure 2.** Scatterplots depicting the relationship between publication year of the studies included in our meta-regression, spanning 2005 to 2023, and two variables: (left) reported accuracy, and (right) total sample size. Pearson’s *r* coefficients and *p*-values are shown in each panel, which represent the monotonic relationship between year and reported accuracy or total sample size.
